# Highly ACE2 Expression in Pancreas May Cause Pancreas Damage After SARS-CoV-2 Infection

**DOI:** 10.1101/2020.02.28.20029181

**Authors:** Furong Liu, Xin Long, Wenbin Zou, Minghao Fang, Wenjuan Wu, Wei Li, Bixiang Zhang, Wanguang Zhang, Xiaoping Chen, Zhanguo Zhang

## Abstract

The ongoing outbreak of coronavirus disease 2019 (COVID-19) caused by severe acute respiratory syndrome coronavirus 2 (SARS-CoV-2) started in the end of 2019 in China has triggered a global public health crisis. Previous studies have shown that SARS-CoV-2 infects cells by binding angiotensin-converting enzyme 2 (ACE2), which is the same as SARS-CoV. The expression and distribution of ACE2 in the pancreas are unknown. At the same time, the injury of pancreas after SARS-CoV-2 infection has not been concerned. Here, we collected public datasets (bulk RNA-seq and single-cell RNA-seq) to indicate the expression and the distribution of ACE2 in pancreas (in both exocrine glands and islets). And further, clinical data including mild and severe patients with COVID-19 demonstrated there existed mild pancreatitis. In the 67 severe cases, 11 patients (16.41%) showed elevated levels of both amylase and lipase, and 5 patients (7.46%) showed imaging alterations. Only one patient (1.85%) showed elevated levels of both amylase and lipase in 54 mild cases, without imaging changes. Our study revealed the phenomenon and possible cause of mild pancreatic injury in patients with COVID-19. This suggests that pancreatitis after SARS-CoV-2 infection should also be paid attention in clinical work.

## Introduction

Severe acute respiratory syndrome coronavirus 2 (SARS-CoV-2), a novel coronavirus that causes coronavirus disease 2019 (COVID-19) in humans, began to occur in Wuhan (China) in December 2019 and has the trend of spreading around the world[1, 2, 3]. By February 28, 2020, a total of 78962 people had been infected in China with 2791 (3.53%) deaths, and 4838 people had been infected with 76 (1.57%) deaths in other countries. It was confirmed by etiology that SARS-CoV-2 belonged to the novel coronavirus of the subgenus Sarbecovirus (Beta-CoV lineage B), and had 79.5% similarity with SARS-CoV[4], which caused the global outbreak in 2003. Spike (S) protein, one of the main structural proteins of SARS-CoV-2, binds angiotensin-converting enzyme 2 (ACE2) protein of the host cell membrane to fuse into the cell for nucleic acid replication just similar to SARS-CoV[5]. Through a variety of bioinformatics and experimental verification, ACE2 is not only expressed in alveolar epithelial cells, but also in the heart, gastrointestinal tract, kidney, testis and other organs, which means that SARS-CoV-2 is likely to enter other tissues and organs through ACE2 binding, causing multiple organ damage[6, 7].

In recent studies, SARS-CoV-2 has been found to cause heart, kidney, liver injury and gastrointestinal symptoms in addition to lung lesions such as ARDS[6]. However, there is lacking attention in pancreatic injury, which may accelerate patients’ progression. In our clinical observation, we also found that some patients showed signs of pancreatic injury, such as high levels of amylase and lipase in serum and urine.

In this study, we used public datasets to explore the expression of ACE2 in the pancreas and various types of pancreatic cells. Combined with clinical data, we demonstrated the pancreatic injury in some patients with COVID-19, and explained for the first time that the pancreas was also a vulnerable organ after SARS-CoV-2 infection.

## Methods

### Analysis of Public datasets

We collected bulk RNA-seq data from GTEx (https://gtexportal.org), which contains normal tissue and organ sequencing data from multiple individuals. The data analysis was performed online. In addition, we collected data from single-cell RNA-seq (scRNA-seq) of the pancreas NCBI-GEO (GSE85241 (4 donors with 2,126 pancreatic cells), GSE84133 (4 donors with 8569 pancreatic cells)). The details of the donors are available in the previous report[8, 9]. The scRNA-seq data processing process is as follows: Unique molecular identified (UMI) expression count matrix was obtained from the database, and Seurat object was created. Further quality control was performed, cells with high mitochondrial gene expression > 5% were filtered. The data was normalized and log-transformed with the method “LogNormalize” in NormalizeData function. Before we performed linear dimensional reduction, the data was scaling. And then principal component analysis (PCA) was performed to the data and determined the dimensionality. Finally, after clustering cells based on graph-based clustering approach, the non-linear dimensional reduction based on uniform manifold approximation and projection (UMAP) was performed the data to analyze and visualize the data. All of scRNA-seq data analysis was based on Seurat R package (version: 3.1.4) with the default parameters[10]. The annotation of cell types was completed based on the featured genes of each cluster and the cell markers of each type pancreas cell from CellMarker database[11] and previously report[8, 9].

### Collections of clinical data

This is a retrospective and observational study from Wuhan Tongji hospital and Wuhan Jin Yin-tan hospital. We retrospectively analyzed patients diagnosed with COVID-19 from January 1, 2020 to February 15, 2020. The criteria for the diagnosis and severity of the patients were followed by the diagnosis and treatment scheme for COVID-19 (trial version 6) issued by National Health Commission of the People’s Republic of China. We collect hospital admissions, laboratory tests, and imaging tests from clinical electronic medical records. In this study, we collected clinical information including age, gender, amylase and lipase in serum, and the imaging results including bedside ultrasound and abdominal CT. Mild COVID-19 patients with serum amylase and lipase in the normal range did not undergo the imaging evaluation of the pancreas, and their pancreas was assumed to be normal. Bedside ultrasound of the abdomen was all performed in critically ill patients, and CT was added if there were any abnormalities.

## Results

To determine the expression of ACE2 in normal pancreas of humans, we used the GTEx database to explore ACE2 expression. We found ACE2 was expressed in many organs or tissues except most brain tissues (Fig. 1A), which suggests that these organs or tissues such as unreported ovary and thyroid might also be targets for SARS-CoV-2. For comparison, we compared ACE2 expression in lung and pancreatic tissues, since the lung is known to be the first target organ to be attacked by SARS-CoV-2. The mRNA level of ACE2 in pancreas was higher than that in lung (Fig. 1A, P < 0.001, Wilcoxon signed rank test).

**Figure 1.**
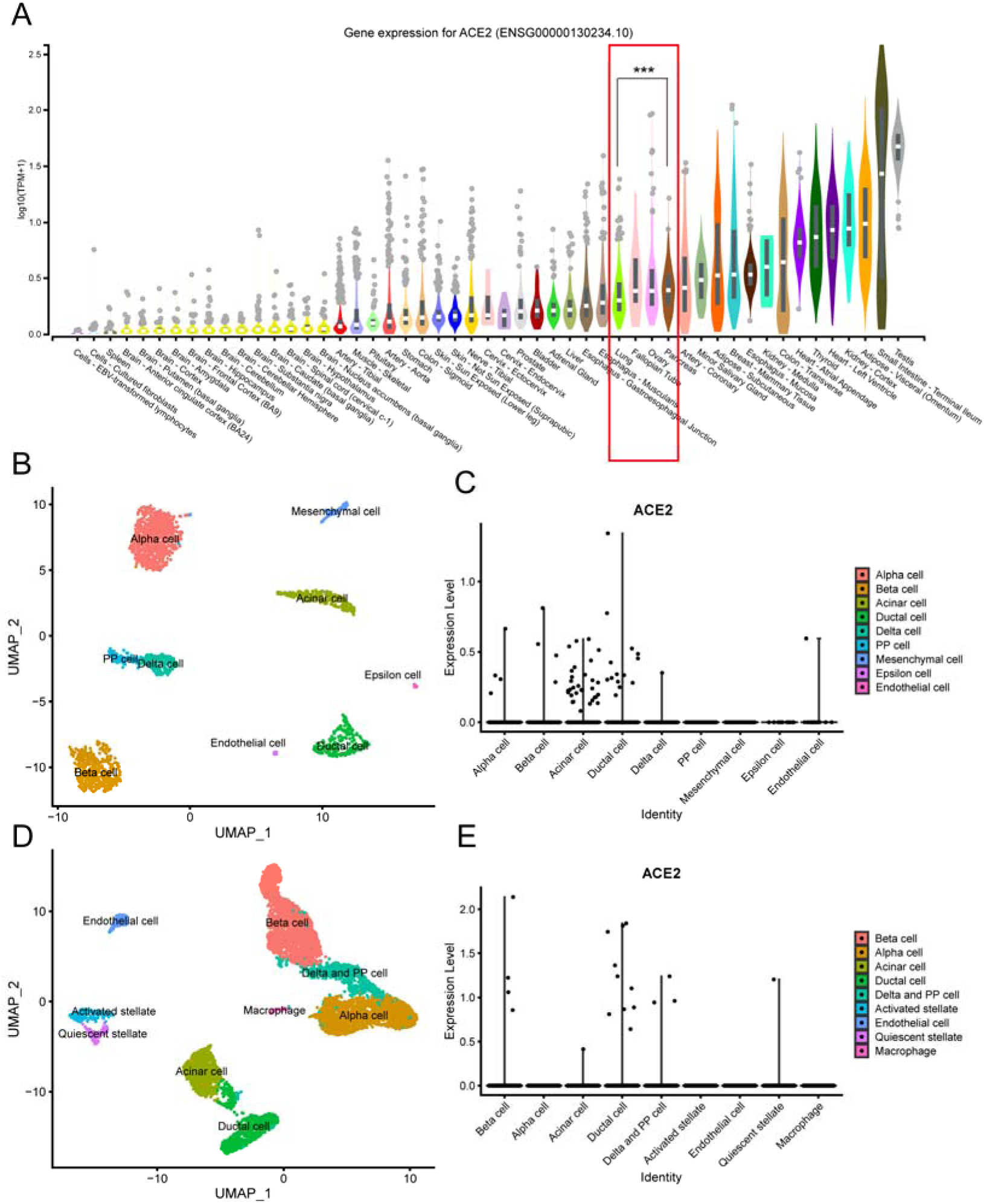
The expression and distribution of ACE2 in pancreas. (A) The mRNA level of ACE2 in multiple organs from GTEx samples. (B), (D) The visualization of pancreatic cell distribution by UMAP in GSE85241 and GSE84133. (C), (E) The expression of ACE2 in different pancreatic cell in GSE85241 and GSE84133. (***: P < 0.001)

To investigate the distribution of ACE2 in the pancreas, we analyzed two scRNA-seq datasets. We identified different types of pancreatic cells, such as alpha cell, beta cell, acinar cell, ductal cell and several other types of cells (Fig. 1B, D). In GSE85241, ACE2 was expressed in 55 cells (2.59%) among the total 2126 cells, most of which (81.82%) were expressed in exocrine gland (duct cells and acinar cells), and a few (16.36%) were expressed in pancreatic islets (alpha, beta, delta and PP cells) (Fig. 1C, Table 1). In GSE84133, 0.22% (19/8550) cells expressed ACE2, with 52.63% (10/19) expressed in duct cells, 5.26% (1/19) expressed in acinar cells, 21.05% (4/19) expressed in beta cells and 15.79% (3/19) expressed in delta and PP cells (Fig. 1E, Table 1). Individual differences between donors from different sources may lead to differences in the analysis results of the two scRNA-seq datasets. These data still indicate that ACE2 is expressed in both the exocrine glands and islets, which may be the main potential part targeted by SARS-CoV-2 in the pancreas, resulting in pancreatic injury.

**Table 1.** The distribution of ACE2 in several type of pancreatic cell in two scRNA-seq datasets.

|  | **GSE85241 (N=55)** | **GSE84133(N=19)** |
| --- | --- | --- |
| **Exocrine gland** |  |  |
| Duct cell | 14 (25.45%) | 10 (52.6%) |
| Acinar cell | 31 (56.36%) | 1 (5.26%) |
| Endothelial cell | 1 (1.82%) | 0 |
| **Pancreatic islet** |  |  |
| Alpha cell | 4 (7.27%) | 0 |
| Beta cell | 4 (7.27%) | 4 (21.05%) |
| PP and delta cell | 1 (1.82%) | 3 (15.79%) |

In our study cohort, among the indicators of pancreatitis we focused on, the main ones were the elevation of serum amylase and lipase. In mild cases, 1.85% (1/54) of patients showed elevated levels of both amylase and lipase, while in severe cases, the proportion of increased amylase was 17.91% (12/64) and the proportion of increased lipase was 16.41% (11/64) (Table 2). However, in the imaging evaluation, only 5 severe patients (7.46%) showed changes in the pancreas, mainly focal enlargement of the pancreas or dilatation of the pancreatic duct, without acute necrosis (Fig. 2). These clinical data show that there exist mild pancreatic lesions in some patients with COVID-19, mainly in severe cases.

**Table 2.** Characteristics of pancreatitis in patients with COVID-19.

|  | **All patients(N=121)** | **Non-severe(N=54)** | **Severe(N=67)** |
| --- | --- | --- | --- |
| **Age (Years)** | 57.3 (18-87) | 52.7 (18-83) | 62.4 (24-87) |
| **Gender** |  |  |  |
| Female | 46 (38.02%) | 21 (38.89%) | 25 (37.31%) |
| Male | 75 (61.98%) | 33 (61.11%) | 42 (62.69%) |
| **Amylase/Lipase increased** | 13 (10.74%) | 1 (1.85%) | 12 (17.91%) |
| Amylase increased | 13 (10.74%) | 1 (1.85%) | 12 (17.91%) |
| Lipase increased | 12 (9.92%) | 1 (1.85%) | 11 (16.41%) |
| **Imaging alteration** |  |  |  |
| A : Normal | 8 (95.86%) | 55 (100%) | 62 (92.54%) |
| B: Enlargement or dilation | 5 (4.13%) | 0 | 5 (7.46%) |
| C: Necrosis | 0 | 0 | 0 |

**Figure 2.**
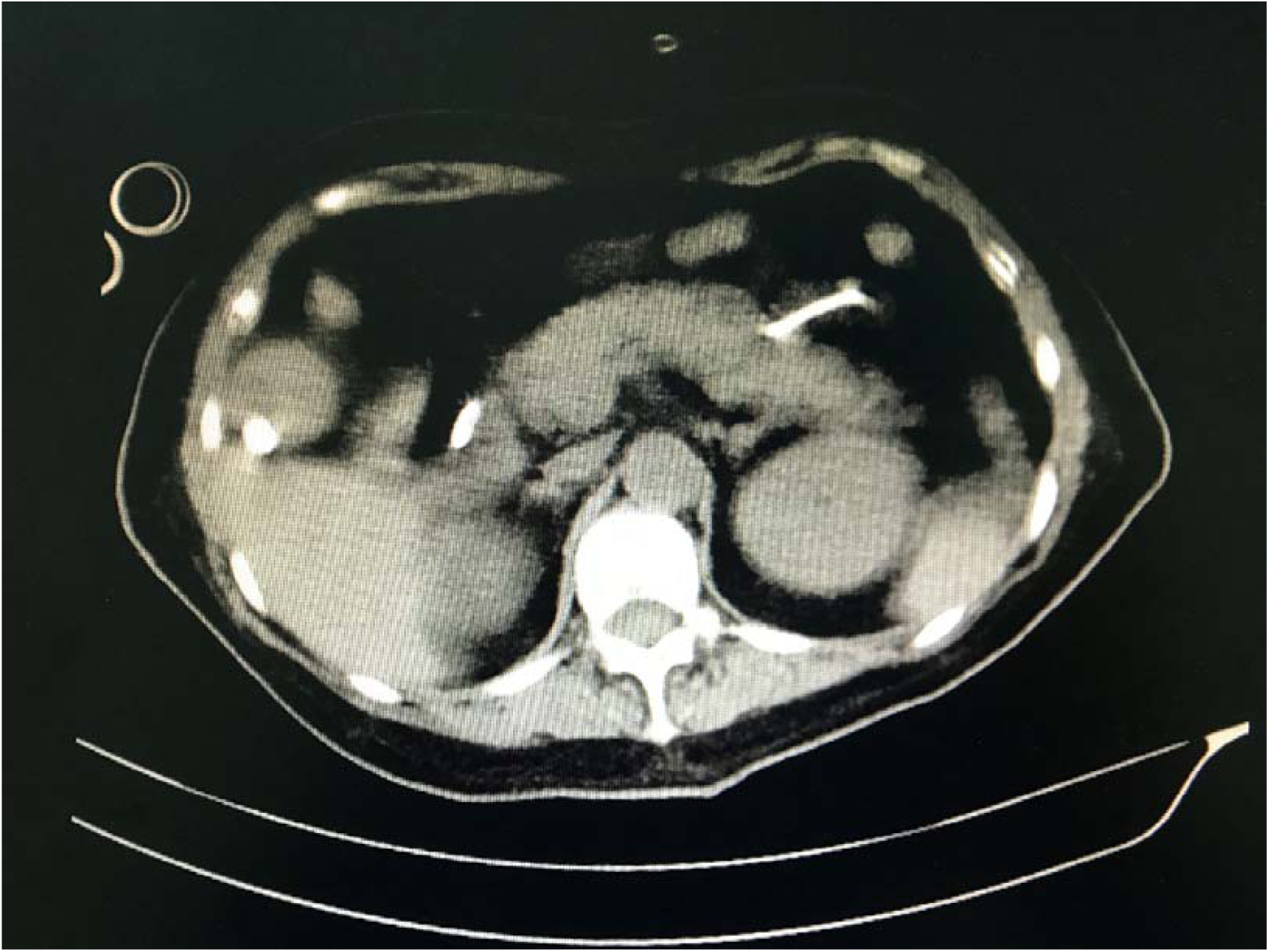
This CT scan shows an enlarged pancreas with dilated pancreatic ducts in a severe patient with COVID-19.

## Discussion

SARS-CoV-2 infection is a very serious public health issue. It caused a nationwide spread in China, and now has the tendency to spread all over the world, causing a surge in infections and deaths[6]. So far, there is no specific drug for SARS-CoV-2, and challenges remain. Although there are asymptomatic infected people, who can also be the source of transmission[1, 12], in clinical practice, patients infected with SARS-CoV-2 mainly suffer from systemic symptoms like fever and fatigue, respiratory symptoms like cough and expectoration, digestive symptoms such as diarrhea[2, 13]. In some patients, especially critically ill cases, damage to other organs, such as the heart, kidneys, and liver, has also been found, which brings great inconvenience to manage patients[2, 14]. As the receptor of SRAS-CoV-2, the expression of protein ACE2 in these organs may provide an approach for SARS-CoV-2 infection[7], leading to tissue injuries. However, the injury of pancreas has been ignored in clinic, since the symptoms of mild pancreatic injury are not specific to other systemic lesions, such as fever, nausea, vomiting and abdominal pain.

In this study, we focused on the expression of ACE2 in pancreas and the damage to the pancreas in a proportion of patients with SARS-CoV-2 infection. Although many previous studies have been reported on the expression of ACE2 tissues and organs[15], including bulk RNA-seq and scRNA-seq data, this analysis in the pancreas is still lacking. Firstly, we found ACE2 was mostly expressed in the pancreas of normal people, and it was slightly higher in the pancreas than in the lungs, indicating that when SARS-CoV-2 appeared in the circulation, it might also combine with ACE2 in the pancreas to enter cells and cause pancreatic injury. Further, we performed scRNA-seq datasets of the pancreas to explore the localized expression of ACE2 in the pancreas. ACE2 is expressed in both exocrine glands and islets. At the same time, analysis results from scRNA-seq data were consistent with elevated levels of amylase and lipase in the plasma of our patients.

In our cases, about 1-2% of the non-severe patients with SARS-CoV-2 had pancreatic lesions, and about 17% of the severe patients suffered from pancreatic injuries, which was not previously noticed. Although their imaging alterations suggested that pancreatitis was not severe, the problem should not be ignored, especially in severe patients. In the clinic, the consequences of pancreatic injury can still be very serious, such as aggravating systemic inflammation, especially in the lungs and accelerating the occurrence of acute respiratory distress syndrome (ARDS)[16], and even developing into chronic pancreatitis without early intervention, which will have a serious impact on the health and quality of life of patients. On the other hand, Yang et al. reported that patients infected with SARS-CoV who had never used glucocorticoids suffered from hyperglycemia, which might be caused by SARS-CoV damaging the pancreatic islets through ACE2[16, 17]. Since SARS-CoV-2 has the same receptor as SARS-COV, whether this result will occur requires more attention. Therefore, increased attention should be paid to the pancreas in patients with SARS-CoV-2 infection, especially in severe cases.

In our study, there are still some limitations. There is no direct evidence that the slight damage to the pancreas was caused by SARS-COV-2 binding to ACE2 in the pancreas, and no studies have shown that nucleic acids of SARS-COV-2 have been detected in the pancreas. Due to the particularity of the location of the pancreas, this study could not be carried out. At present, the body anatomy has been conducted in China, and we hope to see this result in the future. At the same time, the systemic inflammatory response in severe COVID-19 patients might also cause mild damage to the pancreas. However, in the causes of pancreatitis, virus infection factors such as mumps, herpes simplex virus should not be ignored[18]. Our study provides a new idea and a possible mechanism.

## Data Availability

All data generated or analyzed during this study are included in the manuscript.

## Authors’ contributions

FRL, ZGZ conceived this study. ZGZ, BXZ, WGZ, XL, WBZ, MHF, WJW, WL, collected the clinical data. FRL and ZGZ analyzed the data. XPC provided the advices of this study. FRL and ZGZ wrote the manuscript. All authors read and approved the final manuscript.

## Availability of data and materials

All data generated or analyzed during this study are included in the manuscript.

## Ethics approval and consent to participate

This research was authorized by the Ethic Committee of Wuhan Tongji Hospital.

## Consent for publication

All authors agree to submit for consideration for publication in the journal.

